# Atherogenic sdLDL-cholesterol and resistin association with vitamin B12 in relation to Body Mass Index

**DOI:** 10.1101/2024.01.08.24300967

**Authors:** Fauzia Ashfaq, Jwaher Haji Alhaji, Mohammed Salem Alharbi, Fahmida Khatoon, Nayef Hamad E Alhatlani, Fahad Ayash Mari Alkhamisi, Ghareeb O. Alshuwaier, Mohammad Idreesh Khan, Mirza Masroor Ali Beg

## Abstract

**Introduction:** Obesity is a known risk factor for many chronic diseases and is a growing global health concern and poor health outcomes have consistently been linked to body mass index (BMI). Small dense low-density lipoprotein (sdLDL) changes brought on by obesity may increase the likelihood of endothelium penetration and subsequent atherogenesis. Numerous tissues’ metabolic and secretory functions are altered by obesity, which may also increase the serum resistin level.

**Methods:** Present study included 300 participants with diffent BMI among them sdLDL and resistin was evaluated. HbA1c was analysed by whole blood of EDTA and the serum were thawed sample was used for lipid parameters (TG, cholesterol, HDL, LDL, VLDL and sdLDL) and vitamin B12 analysis as well as resistin level was analysed by ELISA.

**Results:** Study observed higher HbA1c (%, p=0.0004), LDL (mg/dl) (<0.0001), TG (mg/dl) (<0.0001), Cholesterol (mg/dl) (<0.0001), VLDL (mg/dl) (<0.0001) in obese compared to overweight, normal BMI, except HDL. Smokers and hypertensive participants had higher sdLDL (p=0.03, p=0.0001) and resistin level (p=0.03, p<0.0001). Obese participants had high amount of sdLDL (p<0.0001, p<0.0001) and resistin level (p<0.0001, p<0.0001) compared to overweight and normal BMI. SdLDL and resistin were found to be positively correlated as well as correlation analysis of sdLDL and resistin level was observed to be significantly correlated with BMI, systolic, TG, cholesterol, VLDL, LDL while negative correlation with HDL level. ROC analysis showed that sdLDL and resistin could be used as prognostic factor for overweight/obesity at cutoff value of 18.55 mg/dl (sdLDL) and 750 pg/ml (resistin). It was also observed that the participant with normal BMI had 389.6 pmol/L while overweight participant had 300.6 pmol/L (p<0.0001) and obese had 291.0 pmol/L (p<0.0001).

**Conclusion:** Study concluded that the obese participants had higher TG, cholesterol, VLDL, LDL and lower HDL level. The most importantly, higher sdLDL level and resistin level was observed in obese participants as well as higher sdLDL and resistin could influence the higher lipid parameters (TG, cholesterol, VLDL, LDL) and lower HDL level. It was also observed that obese participants had lower vitamin B12 level and could lead to other metabolic error.

## Introduction

Body mass index (BMI) has long been associated with worse health outcomes, and many ethnic groups have been made aware of the negative health effects of increase in BMI [1]. BMI is commonly used to assess the level of obesity in therapeutic settings [2] and according to the World Health Organization (WHO), the BMI is divided into three categories: normal (18.5-24.9 kg/m2), overweight (25-29.9 kg/m2), and obesity (30 kg/m2) [3]. It has been discovered that small dense LDL cholesterol (sdLDL-C), an independent CVD risk factor and developing CVD risk biomarker, is a more reliable predictor of CVD [4] and associated with metabolic syndrome [4] as well as obesity or rapid weight gain may raise to be risk of developing diabetes [5], although obesity is not commonly considered to be a diabetes symptom [5]. SdLDL particles are highly linked to metabolic changes that are indicative of insulin resistance, while their primary mechanisms for causing atherogenesis rely on fast buildup inside artery walls and increased oxidation [6]. South Asians are more susceptible to type 2 diabetes and cardiovascular disease due to their central body fat distribution, even at BMIs below the current overweight BMI limit [7]. Patients who are overweight have greater levels of triglycerides, LDL, apolipoprotein glycation, and LDL oxidation sensitivity [8]. Measuring LDL subclasses like sdLDL is especially important to maximize the effectiveness of cardiac disease risk assessment in metabolic syndrome [9]. It has been observed that LDL particle distribution revealed the involvement of sdLDLs in CVD patients and its independent role in the development of CVD [10]. It has been demonstrated that one of the characteristics of obesity-induced dyslipidemia, caused by obesity, is the accumulation of sdLDL particles leads to obesity and insulin resistance could be contributory factor for atherogenic risk [11]. Additionally, it has been established that the most significant predictor of a metabolically unhealthy overweight phenotype is the occurrence of sdLDLs [12]. Obese children have been observed to have a greater occurrence of sdLDL [13], that negatively shift toward more atherogenic particles in childhood and reports that hypertension obese children have higher sdLDL particle [13].

Resistin, an adipokine with a high cysteine content that is produced during adipogenesis, has been linked to insulin resistance in diabetes and obesity [14]. Higher serum resistin levels were found in obese individuals compared to lean individuals [15], and these levels altered in a relation to visceral fat area and BMI [16]. Menzaghi C et al observed that evelated serum resistin levels in non-diabetic, obese people were found to be highly heritable [17]. Resistin levels were observed to be higher in obese individuals compared to individuals with normal body mass [18]. Resistin secretion is interconnected with diabetes, obesity, insulin insensitivity, and adiposity [19], mouse model study revealed that high fat diet or genetic obese mice had elevated resistin level in serum and it has been also observed that the diabetes treatment reduces the serum resistin level [19], Neutralization of resistin or low level of resistin overcome the problem of insulin sensitivity and recovers the glucose concentration [19]. Therefore, present study aimed to understand the alteration in sdLDL level, serum resistin and vitamin B12 level with different body mass index among Saudi Arabian population.

## Materials and Methods

### Subjects and settings

Present study included 300 participants both male and female, categorized on the basis of BMI (normal: 18.5-24.9 kg/m2, overweight: 25-29.9 kg/m2, and obesity: (≥30 kg/m2). Among them 116 (38.7%) were normal body weight, 110 (36.7%) overweight and 74 (24.6) obese respectively. Participants with any chronic underlying disease irrespective of the BMI category were excluded from the study. Informed consent was obtained from the participants before sample collection. 1ml Peripheral blood samples were collected in EDTA vials for HbA1c and 2 ml in plain vials from all the normal weight, overweight, and obese participants. Sample collected in plain vials were centrifuged on 1500 rpm for 10 minutes to separate the serum and stored at −80×C.

### Lipid profile and B12 level Assessment

Sample collected in EDTA vials were subjected for the HbA1c, the serum were thawed for lipid parameters (TG, cholesterol, HDL, LDL, VLDL and sdLDL) and vitamin B12 analysis. Total cholesterol (TC), HDL-C and triglyceride (TG) were measured by an automated enzymatic technique using a Hitachi 917 autoanalyser from Roche diagnostics. LDL-C was calculated using the Friedewald equation [10],LDL-C = TC - (TG/5) - HDL-C. Direct quantitative determination of sdLDL assay was done using LDL-EX ‘‘SEIKEN’’ reagent kits (from Randox laboratories, Limited, Antrim, United Kingdom). vitamin-B12 levels were evaluated using the Cobas e411 electrochemiluminescence immunoassay (Roche, Basel, Switzerland).

### ELISA for resistin

Stored serum in −80×C were thawed and centrifuged and further ELISA for resistin (Human Resistin ELISA Kit, ab183364) was performed using serum from all participants with different BMI (normal weight, overweight and obesity) using 96 well plate following kit protocol (Abcam, San Francisco, CA, US). A vitamin B12 level 148 pmol/L was regarded as insufficient, whereas a level greater than this was seen as normal [20].

### Statistical analysis

The obtained information was recorded in an excel file and analyzed by SPSS.21 and Graph Pad Prism software version 6.05 software. All the nonparametric quantitative data were analyzed by Kruskal–Wallis test (more than 2 group) and Mann–Whitney U-test (2 group). The quantitative parameters were examined for associations using a Spearman correlation test for the variables. The mean±standard deviation was used to express the results and differences between groups were considered significant at P < 0.05.

## Results

### Demographic of study participants

Demographic and general characteristic of study participants were depicted in table 1. Present research work had total 300 study participants among them 85.7% were males and 14.3% were females. Participants with ≤ 40 age group were 61.4% and > 40 years age group were 38.6%.

**Table 1:**
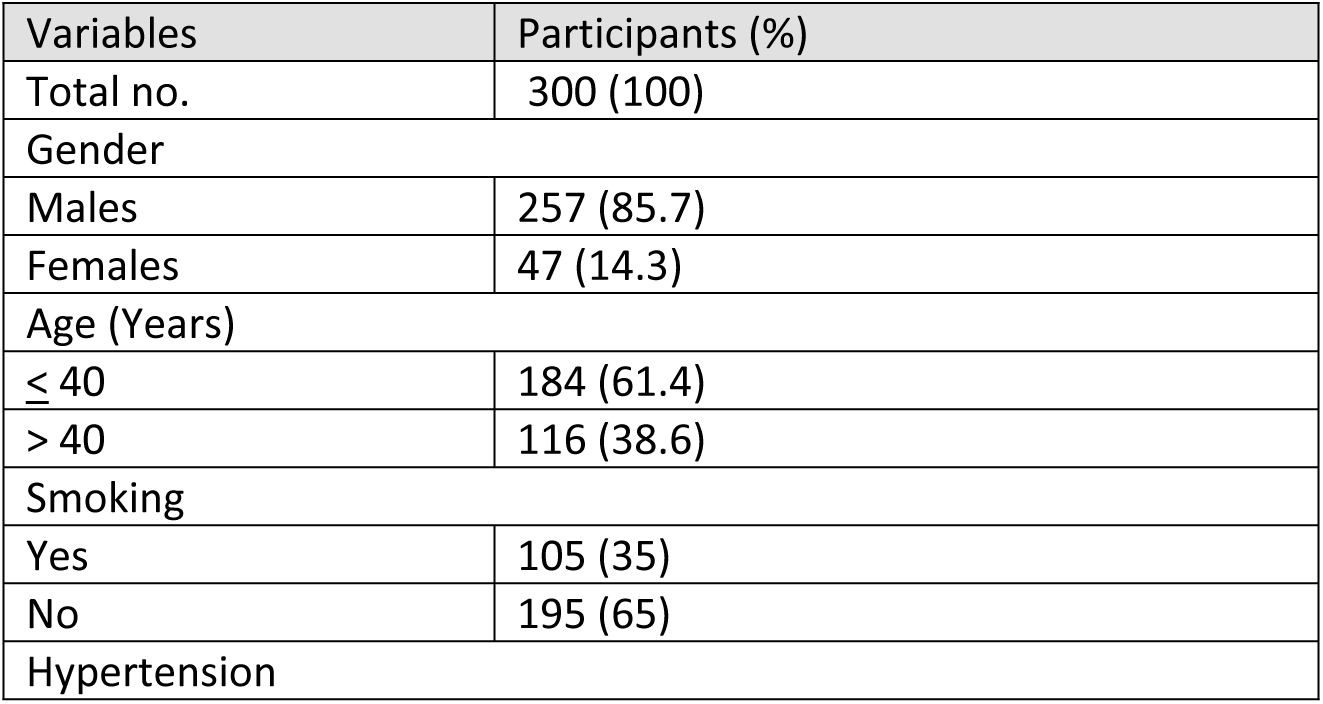
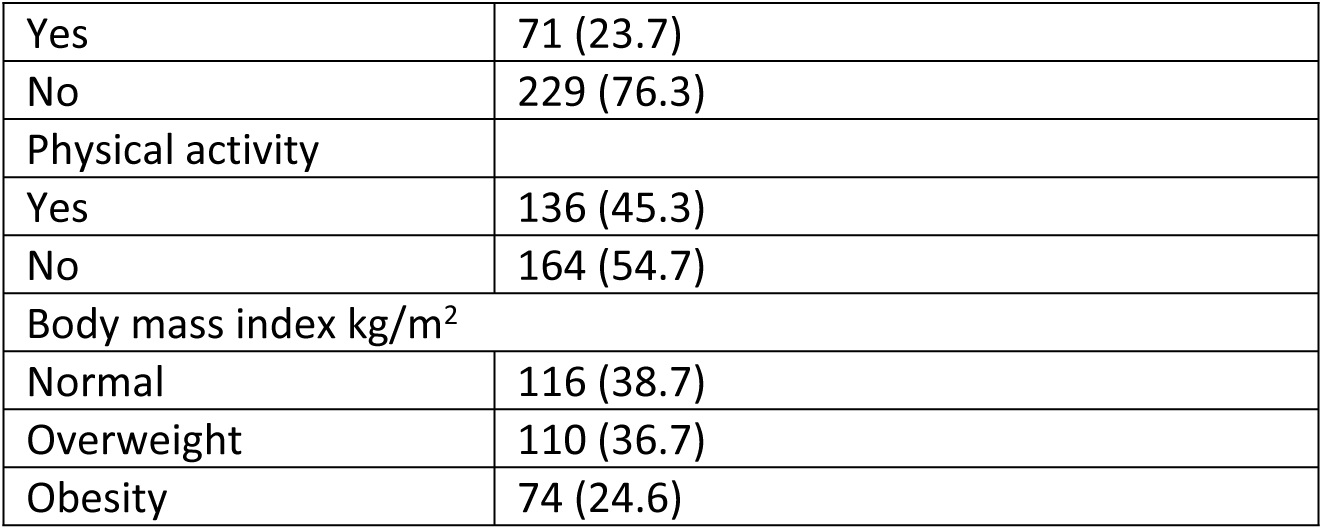
General demographic details of the study’s participants.

| Variables | Participants (%) |
| --- | --- |
| Total no. | 300 (100) |
| Gender |  |
| Males | 257 (85.7) |
| Females | 47 (14.3) |
| Age (Years) |  |
| $\leq 40$ | 184 (61.4) |
| $> 40$ | 116 (38.6) |
| Smoking |  |
| Yes | 105 (35) |
| No | 195 (65) |
| Hypertension |  |
| Yes | 71 (23.7) |
| No | 229 (76.3) |
| Physical activity |  |
| Yes | 136 (45.3) |
| No | 164 (54.7) |
| Body mass index kg/m<sup>2</sup> |  |
| Normal | 116 (38.7) |
| Overweight | 110 (36.7) |
| Obesity | 74 (24.6) |

### Body mass index and biochemical parameters

Biochemical parameters were compared based on the BMI (table 2). HbA1c, LDL, TG, cholesterol, VLDL level was compared between normal BMI, overweight and obese participants and observed to have significant difference. Higher amount of HbA1c (%, p=0.0004), LDL (mg/dl) (<0.0001), TG (mg/dl) (<0.0001), Cholesterol (mg/dl) (<0.0001), VLDL (mg/dl) (<0.0001) was observed among the obese and overweight participants in contrast to normal BMI participants while higher HDL in normal BMI participant compared to overweight and obese participants.

**Table 2:** Clinical and biochemical parameters between healthy controls, overweight participants, and obese participants.

| Variables | Normal BMI | overweight | obese | P value |
| --- | --- | --- | --- | --- |
| HbA1c (%) | 5.07±0.63 | 5.35±0.66 | 5.42±0.65 | 0.0004 |
| HDL (mg/dl) | 44.72±12.82 | 40.42±12.93 | 34.50±6.91 | <0.0001 |
| LDL (mg/dl) | 108.9±19.40 | 187.8±30.60 | 195.3±28.62 | <0.0001 |
| TG (mg/dl) | 164.8±18.02 | 216.9±32.74 | 225.6±39.76 | <0.0001 |
| Cholesterol (mg/dl) | 185.8±32.95 | 233.2±25.30 | 249.1±7.38 | <0.0001 |
| VLDL (mg/dl) | 24.13±5.13 | 32.26±4.51 | 36.92±4.70 | <0.0001 |

### Comparison of sdLDL with different variables

Level of sdLDL was compared with different parameters and clinical outcome of study participants (table 3), it was observed that smoking and hypertension was associated with higher sdLDL among the participant. Participant those who had smoking habit had 21.33mg/dl sdLDL while non-smokers had 19.90 mg/dl (p=0.03). The participants with hypertension history also had higher sdLDL (22.53mg/dl) and non-hypertensive participants had lower sdLDL (19.74mg/dl) comparatively (p<0.0001).

**Table 3:** Comparison of sdLDL level with different variables (gender, age, smoking habit, hypertension and physical activity.

| Variables | sdLDL (mg/dl) | P value |
| --- | --- | --- |
| Gender |  |  |
| Males | 20.47±5.21 | 0.81 |
| Females | 20.11±5.91 |  |
| Age (Years) |  |  |
| ≤ 40 | 20.64±5.04 | 0.84 |
| > 40 | 20.76±5.07 |  |
| Smoking |  |  |
| Yes | 21.33±4.45 | 0.03 |
| No | 19.92±5.67 |  |
| Hypertension |  |  |
| Yes | 22.53±3.73 | 0.0001 |
| No | 19.73±5.56 |  |
| Physical activity |  |  |
| Yes | 20.07±5.50 | 0.29 |
| No | 20.84±5.06 |  |

### Comparison of resistin with different variables

Level of resistin was compared with different parameters and clinical outcome of study participants (table 4) and observed to have statistically significant difference with smoking and hypertension parameters. Participants with smoking habits had higher sdLDL (1047pg/ml) compared to non-smokers (932.2pg/ml) (p=0.01). In the same way for hypertensive participants had 1136pg/ml resistin while non-hypertensive participants had 921.5pg/ml sdLDL (p<0.0001).

**Table 4:**
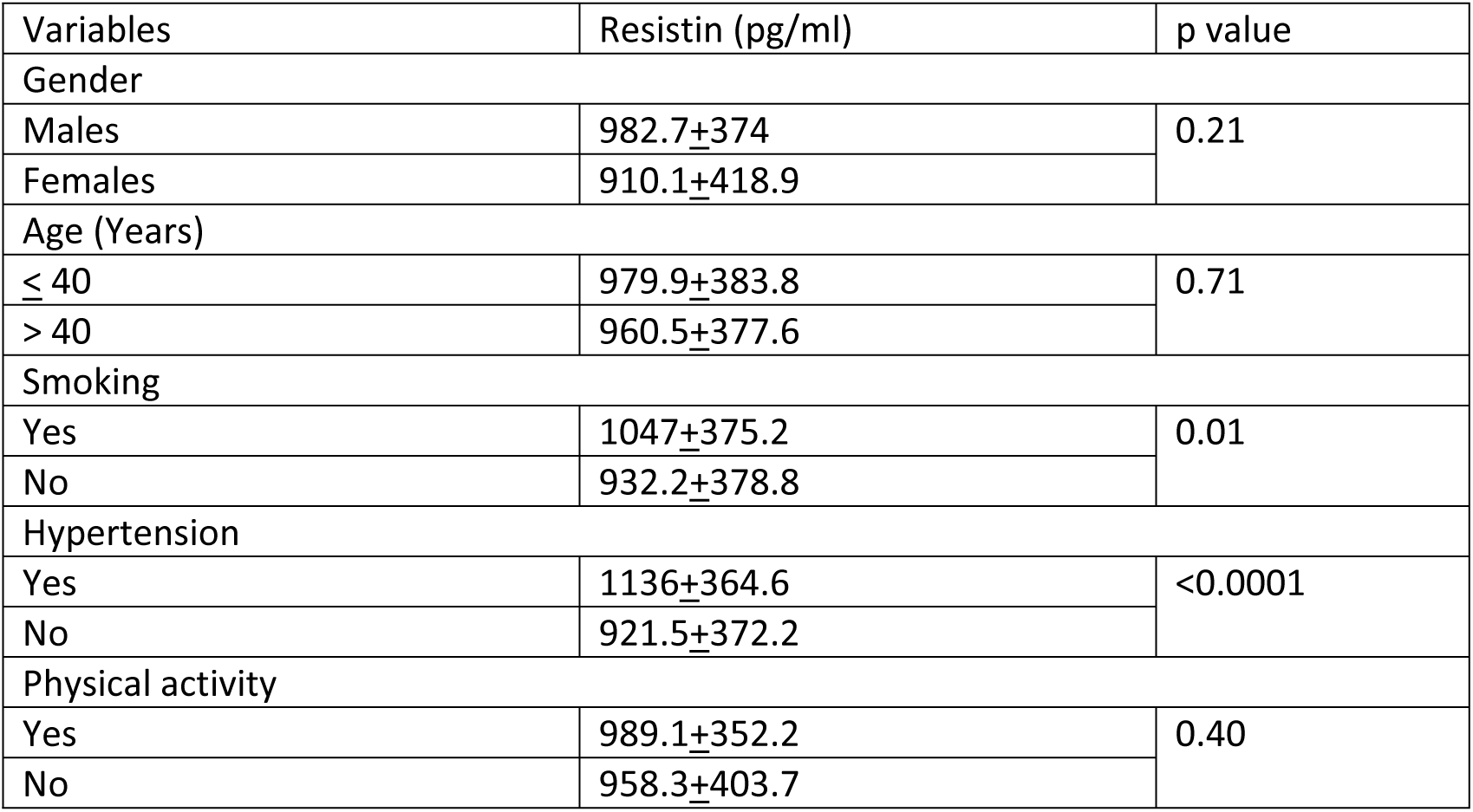
Comparison of resistin level with different variables (gender, age, smoking habit, hypertension and physical activity.

### Association of sdLDL, and resistin with BMI

Present research work compared the level of sdLDL (figure 1a) and resistin (figure 1b) among the study participants with different BMI. It was observed that the normal BMI participants had 15.55mg/dl sdLDL (SD=2.89) while overweight had 21.95mg/dl sdLDL (SD=3.95) (p<0.0001) and obese had 25.76mg/dl sdLDL (SD=3.01) (p<0.0001) level. It was also observed that the normal BMI participants had 603.5pg/ml resistin (SD=163) while overweight had 1003pg/ml resistin (SD=274.9) (p<0.0001) and obese showed 1355pg/ml resistin (SD=220.8) (p<0.0001) level.

**Figure 1:**
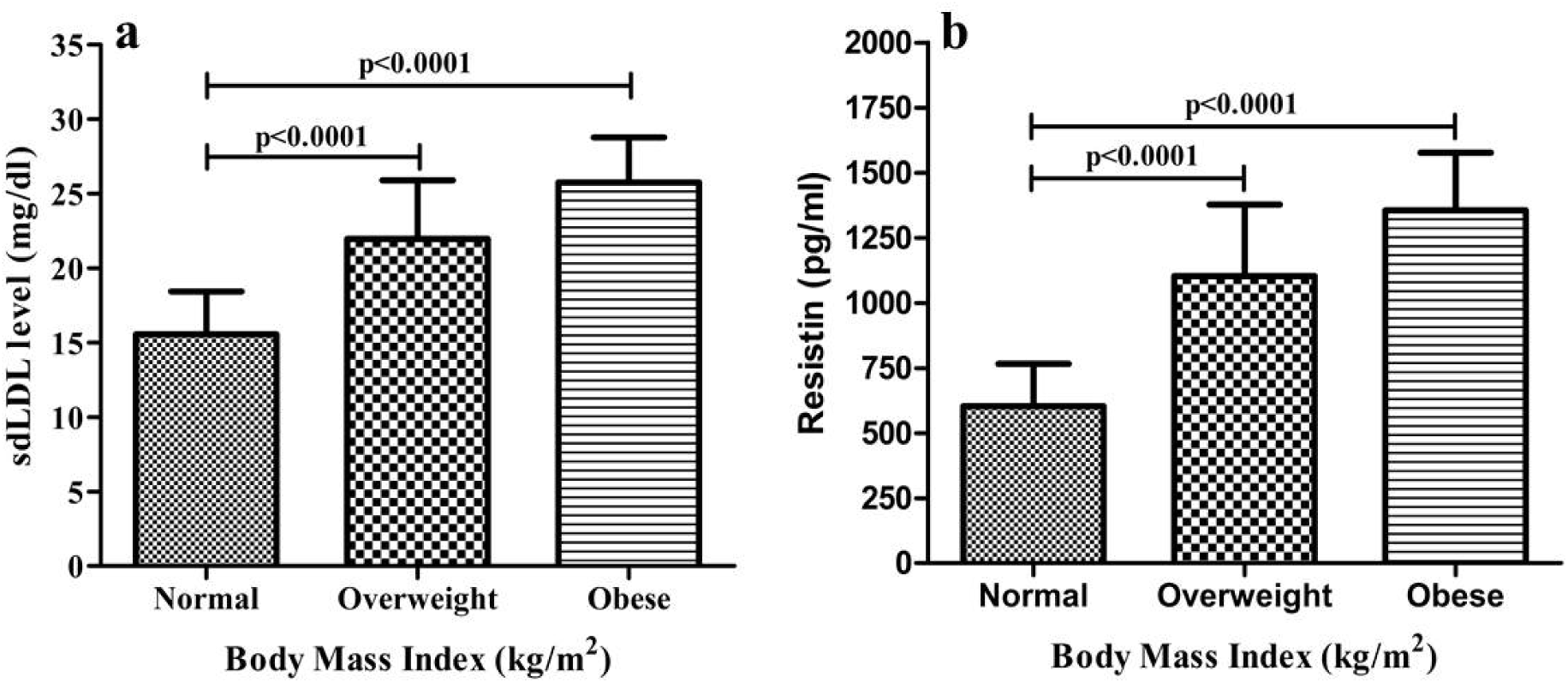
Body mass index, sdLDL and resistin level (a) sdLDL with different BMI (b) Resistin with different BMI.

### Correlation of sdLDL with resistin

A correlation analysis was done between sdLDL and resistin level among the participants to check their association and observed to have positive correlation (r=0.69, p<0.0001). It suggested that increased in sdLDL level could lead to increase in resistin level (figure 2).

**Figure 2:**
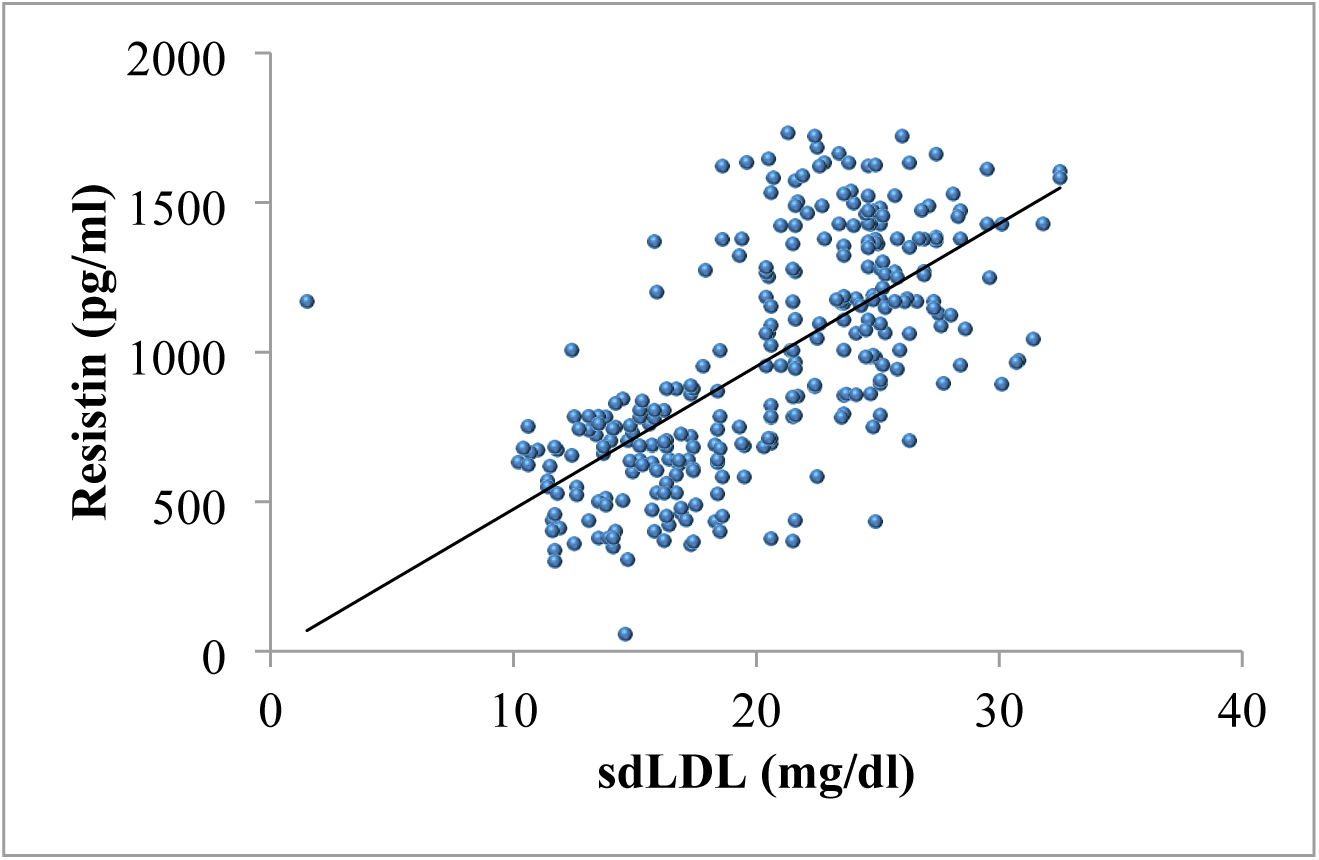
Correlation of sdLDL with resistin level among the study participants.

### Correlation of sdLDL and resistin with clinical parameters

A correlation analysis was done for sdLDL and resistin with clinical/biochemical parameters (table 5). It was observed that sdLDL level was observed to be significantly strongly correlated with BMI (r=0.77), systolic blood pressure (r=0.47), TG (r=0.52), cholesterol (r=0.56), VLDL (r=0.57), LDL (r=0.77) while negative correlation with HDL (r=-0.21) level. In the same way for resistin, it was observed that the resistin level was positively associated with BMI (r=0.81), HbA1c (r=0.15), systolic blood pressure (r=0.50), TG (r=0.60), cholesterol (r=0.61), VLDL (r=0.55), LDL (r=0.63) while negative correlation with HDL (r=-0.29) level.

**Table 5:** Correlation analysis of sdLDL and resistin with clinical parameters such as BMI, HbA1c, systolic, diastolic, HDL, TG, cholesterol, VLDL, LDL.

| sdLDL with clinical parameters | Correlation coefficient (r) | P value | Resistin with clinical parameters | Correlation coefficient (r) | P value |
| --- | --- | --- | --- | --- | --- |
| BMI | 0.77 | <0.0001 | BMI | 0.81 | <0.0001 |
| HbA1c | 0.08 | 0.16 | HbA1c | 0.15 | 0.02 |
| Systolic blood pressure | 0.47 | <0.0001 | Systolic blood pressure | 0.50 | <0.0001 |
| Diastolic blood pressure | 0.09 | 0.10 | Diastolic blood pressure | 0.01 | 0.26 |
| HDL | -0.21 | <0.0001 | HDL | -0.29 | <0.0001 |
| TG | 0.52 | <0.0001 | TG | 0.60 | <0.0001 |
| Cholesterol | 0.56 | <0.0001 | Cholesterol | 0.61 | <0.0001 |
| VLDL | 0.57 | <0.0001 | VLDL | 0.55 | <0.0001 |
| LDL | 0.57 | <0.0001 | LDL | 0.63 | <0.0001 |

### Prognostic significance of sdLDL and resistin

Present study evaluated the prognostic efficacy of sdLDL and resistin (table 6). ROC curve was plotted between normal BMI vs overweight and obesity by plotting for sdLDL, normal BMI was taken as reference vs overweight and obesity, The observed AUC was 0.94, sensitivity 90%, specificity 88% with cutoff value of 18.55mg/dl (figure 3a, p<0.0001). For resistin ROC, the AUC was 0.96, sensitivity 93%, specificity 81% with cutoff value of 750pg/ml (figure 3b, p<0.0001).

**Figure 3:**
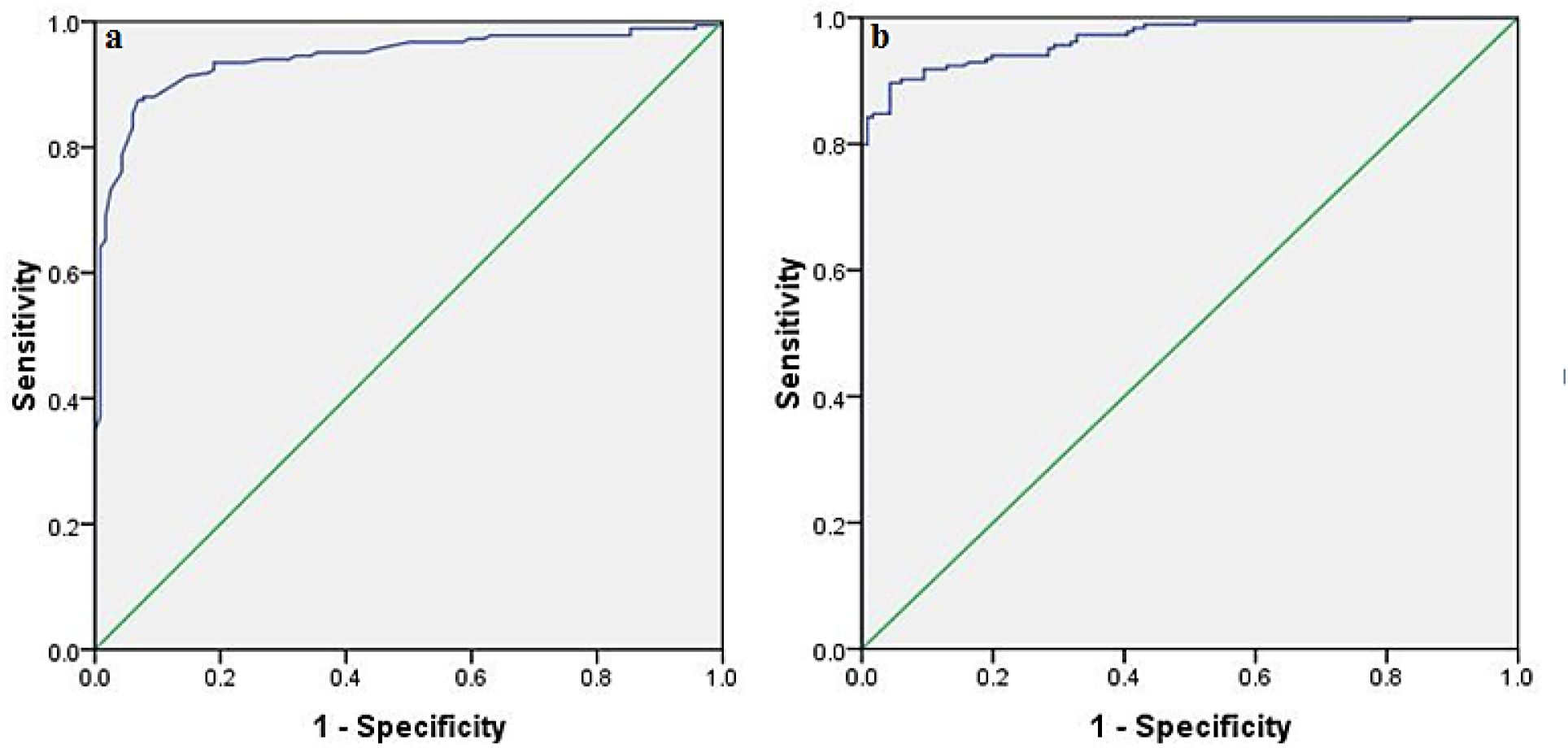
ROC for normal BMI vs overweight and obesity (1) sdLDL for normal BMI vs overweight and obesity (b) resistin for normal BMI vs overweight and obesity.

**Table 6:** ROC curve for sdLDL and resistin for their prognostic efficacy.

| Parameters | AUC (95% CI) | Sensitivity | Specificity | Cutoff | P value |
| --- | --- | --- | --- | --- | --- |
| sdLDL | 0.94 (0.91-0.96) | 90% | 88% | 18.55 mg/dl | <0.0001 |
| Resistin | 0.96 (0.95-0.98) | 93% | 81% | 750 pg/ml | <0.0001 |

### BMI and vitamin B12

Vitamin B12 level was analyzed with respect to BMI (normal vs overweight and normal vs obese) (figure 4). It was observed that the participant with normal BMI had 389.6pmol/L (SD=121.2) vitamin B12 while overweight participant had 300.6 pmol/L (SD=89.5) vitamin B12 (p<0.0001) and obese had 291.0pmol/L vitamin B12 (SD=44.6) (p<0.0001).

**Figure 4:**
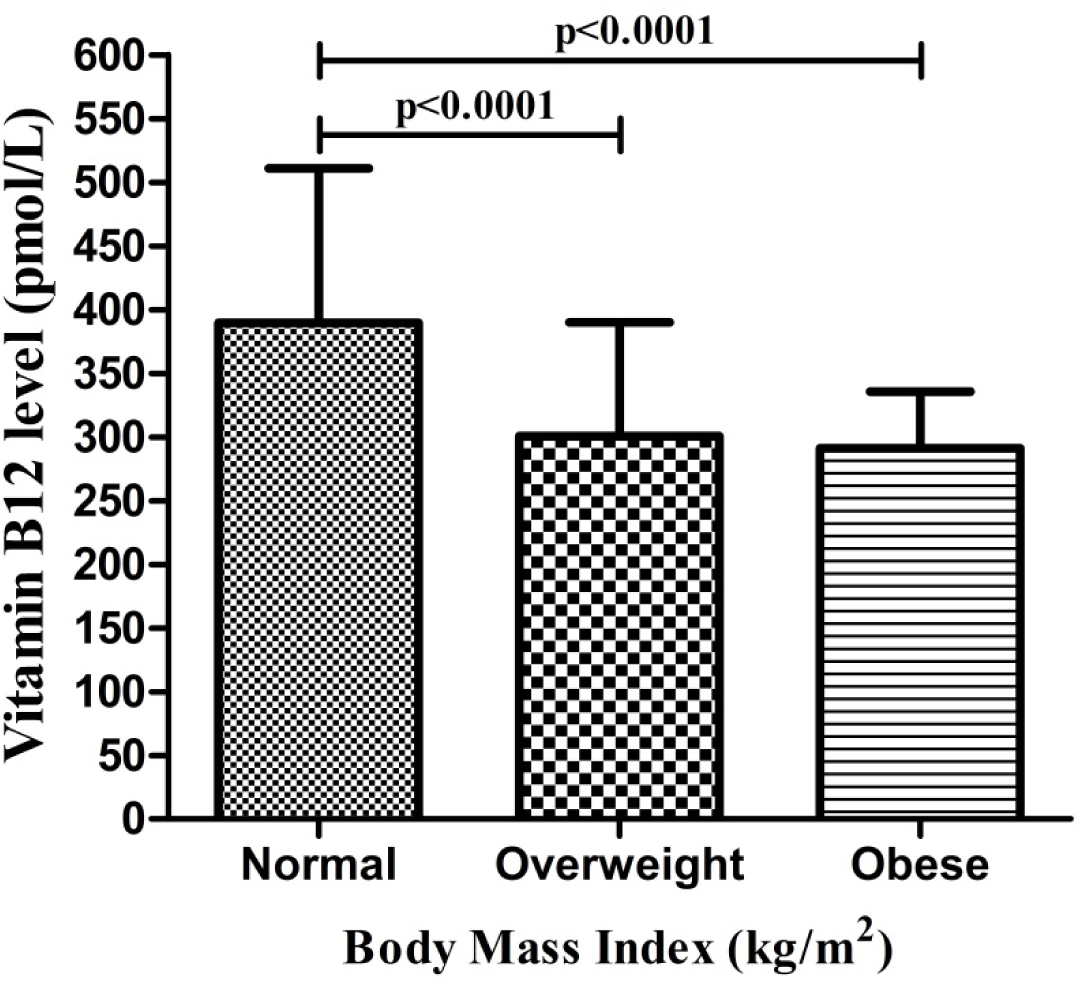
Estimation of vitamin B12 level with respect to BMI (kg/m^2^).

## Discussion

Obesity and metabolic syndrome are both public health issues that are estimated to affect more than one billion persons globally [21]. There has been a substantial increase in global obesity, which has led to an increase in human morbidity and death. Morbid obesity is considered a serious health risk [22]. As previously noted that metabolic consequences of obesity in people are linked to higher risks for a number of illnesses, including insulin resistance, cardiovascular disease, hypertension, and several types of cancer [21]. Less than 25% of male teenagers are sufficiently active globally [23], and in adulthood, this percentage decreases even more [24]. High BMI is also a growing concern in East and Southeast Asia, where the prevalence of obesity has increased quickly in young males [25]. SdLDL concentration is connected with a higher susceptibility for endothelial penetration and subsequent atherogenesis [26] and several biomarkers, including elevated blood TG, TC, and LDL, are linked to elevated sdLDL [27]. High sdLDL levels are influenced by a number of illnesses, including obesity [26], MetS, [11], systemic hypertension [28], and hepatic conditions [29].

Present research work, biochemical parameters were compared between normal BMI, overweight and obesity, higher HbA1c, LDL, TG, Cholesterol, VLDL was observed in obese and overweight participants while lower HDL level compared to normal BMI. Importantly, we observed higher sdLDL level among obese (BMI: ≥30 kg/m2) participant compared to overweight (BMI: 25-29.9 kg/m2) and normal (BMI: 18.5-24.9 kg/m2) BMI participants. We also observed higher sdLDL in hypertensive, smokers compared to non-hypertensive and non-smokers. In the same way Nakamura M et al in 2021, found that smoking cigarettes raises sdLDL levels and that sdLDL levels were significantly lower in people who had done smoking for more than five years [30]. SdLDL, known to be raised in obese people, has been found to have a significant atherogenicity and, enhanced risk of atherosclerotic cardiovascular diseases [26]. Adaja TM. et al., suggested that substantial variation in the level of sdLDL across the three categories of body mass index, and observed highest levels of sdLDL among obese participants [31]. Serum sdLDL concentration showed to be higher among the obese subjects compared to normal weight subjects [32], as well as plasma sdLDL concentration was also high in obese participants [33]. Participants with elevated sdLDL concentrations are thought to have a roughly three- to seven-fold increased risk of developing coronary heart disease [34]. Jiahua Fan et al in 2019, higher concentration of sdLDL are linked to metabolic syndrome (MetS), central adiposity and inflammation [35]. Cross-sectional and prospective observational have shown that people with higher sdLDL levels have a higher risk of developing cardiovascular disease [11]. According to Nikolic D et al in 2013, systemic inflammation and high levels of circulating sdLDL were also linked to obesity [36]. In addition to a study by Sirikul K et al., in 2015, they also discovered that participants who were obese had higher plasma sdLDL particles [37]. According to Bamba V et al in 2007, weight gain has been associated with an atherogenic lipid profile, which is defined by elevated levels of TG, LDL, and sdLDL in the bloodstream and lower levels of HDL [38]. In the same way we also observed elevated TG, LDL and sdLDL among the obese participants.

It was observed that the obese (BMI: ≥30 kg/m2) and overweight (BMI: 25-29.9 kg/m2) participants had higher resistin level in contrast to normal BMI. Resistin showed to play important roles in the pathophysiology of obesity and to linked with atherosclerosis, inflammation, and obesity [39]. It has been observed that the patients with insulin resistance had high resistin concentrations and showed to be higher in obese diabetic patients [40]. It has been observed that resistin concentration was shown to be higher in people with heart failure, left ventricular dysfunction, and CVD [41]. According to a recent study, obese patients had significantly higher resistin levels than participants with normal body mass [42]. Elevated resistin level was observed in obese diabetic women compared to both over-weight and lean patients [43]. Recent human study showed higher resistin level in obese subjects compared to lean subjects suggested that to be associated with obesity [44]. The pathologic development of human obesity is influenced by a variety of factors, particularly lifestyle [45]. Obesity alters the metabolic and secretory activity of numerous tissues [46] and has been linked to elevated serum levels of resistin in humans [47]. A high blood resistin level has been linked to the development of insulin resistance, type 2 diabetes, atherosclerosis, and cardiovascular diseases in animal and human study [48]. Serum resistin levels in obese participants were significantly greater than normal weight subjects, linked with altered TG and adversely with HDL level [49], which is also consistent with present study findings.

Obese study participants with hypertension also had greater serum resistin levels, indicating that resistin was observed to be cause of hypertension [50]. According to Guesoy G et al, plasma resistin level was found to be linked with smoking in turkish population [51]. In the same way present study also observed higher resistin level in hypertensive participants compared to non-hypertensive participants as well as in smokers. A positive correlation was found between sdLDL and resistin in study participants suggested that increase in sdLDL could lead to increase in resistin level. SdLDL and resistin was observed to be positively correlated with BMI, HbA1c, systolic, diastolic, TG, cholesterol, VLDL, LDL and negatively with HDL level. Prognostic significance was also calculated and revealed that the sdLDL and resistin level could be the prognostic factor to predict the condition of overweight and obesity at the cutoff value of 18.55mg/dl for sdLDL and 750pg/ml for resistin. Which could help to control reduce the chance of having other related comorbidity. Low vitamin B12 level was also linked and poor metabolic health profiles, obesity, insulin resistance, and cardiovascular issues [52]. Lower serum vitamin B12 levels have been linked to high BMI, and altered metabolic health, including obesity in Danish communities [53]. According to a 2006 study by Pinhas-Hamiel O et al, lower vitamin B12 level is more prevalent in obese people, including obesity in childrens and adolescents [54] as well as negative correlation between serum vitamin B12 levels and obesity [55–57]. Similar to this, we also noticed lower vitamin B12 levels in obese subjects compared to overweight and normal BMI participants.

## Conclusion

Present study concluded that the obese participants had higher TG, cholesterol, VLDL, LDL and lower HDL level. The most importantly, higher sdLDL level and resistin level was observed in obese (BMI: ≥30 kg/m2) and correlation analysis suggested that higher in sdLDL and restin could influence the higher lipid parameters (TG, cholesterol, VLDL, LDL) and lower HDL level. Higher sdLDL and resistin observed to be the important prognostic marker for higher BMI and could lead to other related comorbidity. It was also observed that obese participants had lower vitamin B12 level and could lead to other metabolic error.

## Data Sharing Statement

We confirm that the data used during the research will not be shared with anybody/broadcasted in any public domain. The datasets generated during and/or analysed during the current study are available from the corresponding author on reasonable request.

## Ethics and Informed Consent

The research ethics committee University of Hail and Ministry of health ethics committee gave their approval to this study. Before the study began, participants gave their written informed consent. The study was carried out in accordance with the Declaration of Helsinki and all ethical principles concerning human experimentation were followed.

## Acknowledgment

The author thanks to researchers supporting project number (RSPD2023R1013), King Saud University, Riyadh, Said Arabia.

